# Results publications are inadequately linked to trial registrations: An automated pipeline and evaluation of German university medical centers

**DOI:** 10.1101/2021.08.23.21262478

**Authors:** Maia Salholz-Hillel, Daniel Strech, Benjamin Gregory Carlisle

## Abstract

**Objective:** To evaluate links between registration and publication across clinical trials led by German university medical centers (UMCs) and registered in either ClinicalTrials.gov or the German Clinical Trials Registry (DRKS). Inadequate links make trial publications and registrations less findable and compromise evidence synthesis and health policy decision making. The World Health Organization (WHO) and others call for better adoption of this straightforward transparency practice.

**Design:** Cross-sectional bibliographic study

**Setting:** German UMC clinical trials

**Methods:** We used automated strategies to download and extract data from trial registries, PubMed, and trial publications for a cohort of all registered, published clinical trials conducted across German UMCs and completed between 2009 and 2017. We implemented regular expressions to detect and classify publication identifiers (DOI and PMID) in registrations, and trial registrations numbers (TRNs) in publication metadata, abstract, and full-text.

**Main outcome measures:** The proportion of trial registrations that reference a known results publication. The proportion of results publications that report the known TRN in the metadata, abstract, and full-text.

**Secondary analyses:** We constructed exploratory logistic regression models to investigate the relationship between trial completion date, registry, and registration-publication linking.

**Results:** Only 20% (373/1,895) of trials have a linked publication (DOI or PMID) in the registration as well as the TRN in the publication full-text, abstract, and metadata, and only 25% (477) of trials met the CONSORT and ICMJE guidelines to include TRNs in both the full-text and the abstract. 17% (327) of trials had no links. The most common link was TRN reported in the full-text (60%, 1,137). ClinicalTrials.gov trials were overall better linked than DRKS trials, and this difference appears to be driven by PubMed and registry infrastructure, rather than by trialists. Of trials reporting a TRN in the abstract, trials in ClinicalTrials.gov were more likely than trials in DRKS to have the TRN captured in the PubMed metadata. Most (78%, 662/849) ClinicalTrials.gov registrations with a publication link were automatically indexed from PubMed metadata, which is not possible in DRKS.

**Conclusion:** German UMCs have not comprehensively linked trial registrations and publications by both including a structured reference to the publication in the registration, and reporting TRNs in results publications. In addition to improved linking by trialists, changes in the PubMed TRN capturing process (such as automated strategies like those developed in this study) and automated indexing of publications in DRKS would make trial registrations and results more findable.

**Open Data and Code:** All code and the final analysis data for this study are available at https://github.com/maia-sh/reg-pub-link. Raw data is available at https://github.com/maia-sh/intovalue-data.

## 1 Introduction

Linking trial registrations and results publications makes each more findable and improves research transparency at minimal effort to researchers. Threaded evidence also empowers readers to cross-check sources, e.g., for potential outcome switching (Altman, Furberg, Grimshaw, & Shanahan, 2014). If trial results cannot be found or linked to their registration, systematic reviewers may miss relevant data and draw invalid conclusions (Miron, Gonçalves, & Musen, 2020). Health policy decisions and clinical guidelines rely on such evidence synthesis, and incomplete evidence can misinform subsequent clinical trials, drive the misallocation of healthcare resources, and risk patient wellbeing (Chan et al., 2014; Council for International Organizations of Medical Sciences & World Health Organization, 2016).

To inform clinical decision making with comprehensive trial evidence, legal regulations and ethical guidelines advocate results transparency (Altman & Moher, 2014; Borysowski, Wnukiewicz-Kozłowska, & Górski, 2020; Chalmers, Glasziou, & Godlee, 2013; World Health Organization, 2017; World Medical Association, 2013). Trial results should be bidirectionally linked, meaning both a reference to publications in the registration, and trial registration numbers (TRNs) in the full-text, abstract, and metadata of publications. The Consolidated Standards of Reporting Trials (CONSORT) as well as the International Committee of Medical Journal Editors (ICMJE) ask trialists to report the “trial registration number and name of the trial register” in both the full-text and abstract of trial results publications (Hopewell, Clarke, Moher, Wager, Middleton, Altman, Schulz, & Group, 2008; International Committee of Medical Journal Editors (ICMJE), 2019). Reporting a TRN solely in the full-text does not suffice, since readers may not have access to the full-text or may judge a trial on the abstract alone (Hopewell, Clarke, Moher, Wager, Middleton, Altman, & Schulz, 2008). Abstracts may also be published independently of a full-text publication, such as for conferences (Hopewell, Clarke, Moher, Wager, Middleton, Altman, Schulz, & Group, 2008).

The inclusion of TRNs in publication and bibliographic metadata especially enhances machine readability and discoverability. Trial results can then be found more efficiently using TRNs to search publication databases (Huser & Cimino, 2013b). Publications can then also be automatically linked within trial registrations, as is currently done with ClinicalTrials.gov using TRN metadata from PubMed (National Library of Medicine, 2017). Such references to results publications within a registration allow readers to quickly identify and navigate to the trial findings. Results references may also be entered manually in selected registries, with varying degrees of structure (Miron et al., 2020; Moja et al., 2009; Venugopal & Saberwal, 2021). ClinicalTrials.gov, for example, provides fields for DOI and PubMed identifier (PMID), whereas the German Clinical Trials Registry (DRKS) offers an unstructured free-text field. While registry readers may be able to find the referenced publication from the free-text, unique identifiers provide a more robust and structured link that allows for automated retrieval (Huser & Cimino, 2013b).

Trial evidence appears to be insufficiently linked in both registrations and publications. Previous studies on trial registrations found links to results publications in as few as 13% to as many as 60% of trials (Bashir, Bourgeois, & Dunn, 2017; Huser & Cimino, 2013b; Wetering, Scholten, Haring, Clarke, & Hooft, 2012). TRN reporting in either the metadata, abstract, or full-text of trial publications has been found to be as low at 8% and as high as 97%, varying widely depending on how the trial population was defined (Al-Durra, Nolan, Seto, & Cafazzo, 2020; Bashir et al., 2017; Carlisle, 2020; Huser & Cimino, 2013a; Wetering et al., 2012). The sampling strategies used in previous studies limit our ability to draw conclusions about registration-publication linking. Publication-based cohorts relied on inaccurate PubMed clinical trial filters (Glanville, Kotas, Featherstone, & Dooley, 2020) and included publications beyond trial results, as well as trials with unknown registration rates, as not all trials are registered, (Denneny, Bourne, & Kolstoe, 2019; Gopal et al., 2018; Trinquart, Dunn, & Bourgeois, 2018). Registration-based cohorts included trials that may not have an associated publication and thus should not be expected to have a publication link (Chen et al., 2016; Dwan, Gamble, Williamson, & Kirkham, 2013; Ross et al., 2012). Furthermore, these studies of publication links in registrations have primarily focused on ClinicalTrials.gov, and we are not aware of previous studies that investigate publication links in DRKS. In this study, we limit our sample to registered clinical trials with published results which can be expected to link the registration and publication, and which allows us to draw conclusions about the prevalence of this responsible research practice.

### 1.1 Objectives

In this exploratory study, we evaluate the links between registrations and results publications in a cross-section of published clinical trials led by German university medical centers (UMCs) and registered in either ClinicalTrials.gov or DRKS with completion dates between 2009 and 2017. We also looked at the relationship between different link types, as well as registry and change in practices over time. We developed an automated and scalable approach for data collection and extraction using regular expressions to detect and classify publication identifiers and TRNs, which may be applied to other trial cohorts.

## 2 Methods

### 2.1 Automated pipeline for data collection

We used two cohorts of registered clinical trials and associated results previously developed by Wieschowski et al. (2019) and Riedel et al. (2021b) and referred to as the “IntoValue” dataset (Riedel et al., 2021a). The dataset consists of clinical trials registered on ClinicalTrials.gov or DRKS, conducted at a German UMC, and completed between 2009 and 2017. Corresponding results publications were found via manual searches.

We downloaded data from both registries on 15 August 2021. We queried ClinicalTrials.gov using the Clinical Trials Transformation Initiative (CTTI) Aggregate Content of ClinicalTrials.gov (AACT) via its PostgreSQL database API (Clinical Trials Transformation Initiative (CTTI), n.d.). As DRKS does not provide an API, we built a webscraper to capture the necessary fields (Federal Institute für Drugs and Medical Devices (BfArM), n.d.).

We queried the PubMed Entrez Programming Utilities (E-utilities) API on 15 August 2021 for all trial results PMIDs (U.S. National Library of Medicine, n.d.; Winter, 2017). From the PubMed XML, we extracted bibliometric information, including the publication abstract and secondary identifier (or databank) metadata.

We used DOIs and PMIDs to search for full-text publications as PDFs, using a combination of automated and manual strategies, including contacting the corresponding author as a final step (Jahn, 2021; Our Research, n.d.). We then used the GROBID machine learning library for technical and scientific publications (v0.6.1) via Python to parse the PDFs into machine-readable XMLs (“GROBID,” 2008; Lopez, 2021). In order to isolate the main body from the publication abstract for our analysis, we extracted the <body> sections of the papers.

#### 2.1.1 Inclusion and exclusion criteria

After updating registry data, we reapplied the IntoValue exclusion criteria: completion date before 2009 or after 2017, non-interventional, incomplete based on study status, non-German-UMC lead. See Riedel et al. (2021b) for further details on these criteria. We limited our sample to trials with results with a PMID and full-text publication.

### 2.2 Detection and classification of publication identifiers in registrations

As each registry formats references to publications differently, we developed parallel approaches to extract publication identifiers for each. For ClinicalTrials.gov, we retrieved the reference type, citation, and PMID fields from AACT. We then used a regular expression (regex) to extract any DOIs from the citations. For DRKS, we scraped the reference type, citation, and URL, when available. We used regexes to extract any DOIs and PMIDs from the citation and URL. In the rare cases when conflicting DOIs or PMIDs were found in the citation vs. the URL, we manually reviewed to determine which was valid; if both were valid, we preferred the identifier provided in the citation. In addition, we determined whether a reference was manually or automatically indexed in the registry. DRKS allows only manually added references; for ClinicalTrials.gov, references of type “derived” were marked as automatically indexed. We then used DOIs and PMIDs to match the referenced publications to those in our trial dataset. We did not attempt to match publications without identifiers (i.e., publication title only).

### 2.3 Detection and classification of trial registration numbers in publications

We developed regexes for the TRN patterns for all PubMed-indexed and ICTRP-network registries (available at https://github.com/maia-sh/ctregistries/blob/master/inst/extdata/registries.csv) and used these to detect and classify TRNs in the PubMed secondary identifier metadata, abstract, and full-text. To gauge the sensitivity and specificity of these regexes, we visually inspected all PubMed secondary identifier metadata. For each source, we first extracted all unique TRN patterns within a source. Our regex allowed for minor formatting errors in the TRN patterns (such as additional punctuation). We then cleaned the TRN patterns to remove these formatting errors, and then deduplicated the corrected TRNs to exclude duplicates within a source uncovered through the cleaning process. We also merged our sources to produce a list of unique TRNs by publication along with the source(s) in which each was reported.

The output of TRN extraction, cleaning, and deduplication included TRNs beyond those of the known registrations. These additional TRNs could be cross-registrations of the same trial or provided as background or discussion. For this study, we were interested in whether the known registrations were reported in the publication.

### 2.4 Analysis

We generated descriptive statistics on trial and publication characteristics, overall and by registry. We calculated the number and proportion of trials linked via the publication (full-text, abstract, metadata) and the registration. To explore change over time, the relationships between types of links, and differences between registries, we ran logistic regressions for each link type, with all other link types as well as registry and completion year as explanatory variables. Regressions were performed as exploratory analyses, and all variables were included in each regression model. There was no model selection or fitting, or correction for multiple testing. In particular, since PubMed metadata may be generated from the publication abstract or full-text, we examined the relationship between TRN reporting in either the abstract or full-text, and TRN inclusion in the metadata. Additionally, since ClinicalTrials.gov registrations automatically reference publications with the TRN in the PubMed metadata (whereas DRKS does not), we examined the proportion of automatically vs. manually linked publications in ClinicalTrials.gov. To explore registry differences for only manually linked publications, we excluded automated links and calculated the number and proportion of trials with the publication linked in the registration.

### 2.5 Software, code, and data

Data collection, preparation, and analysis was performed in R [Version 4.1.0; R Core Team (2021)]. All code, the final analysis data, and a list of all packages used for this study are available at https://github.com/maia-sh/reg-pub-link. Raw data (with the exception of the full-text of publications) is available at https://github.com/maia-sh/intovalue-data.

## 3 Results

The IntoValue dataset includes all trials led by a German UMC that were registered on ClinicalTrials.gov or DRKS and completed between 2009 and 2017 (n = 3790). After applying our exclusion criteria, our sample included 1895 trials with 1861 unique results publications indexed in PubMed and available as full-text, as some publications include results from more than one trial. Appendix A provides a flow diagram of the trial and publication screening. Table 1 shows summary descriptive details of the trials with results publications by registry.

**Table 1.** Characteristics of German UMC-led trials with published results. A trial was considered randomized if allocation included randomization. A trial was considered prospectively registered if registered in the same or previous months to start date. Summary results were taken from a structured data field in ClinicalTrials.gov, and determined based on manual inspection for terms such as *Ergebnisbericht* or *Abschlussbericht* in DRKS. Top journals refer to the journals with the greatest number of trial publications in our sample.

|  | Overall<br>N = 1,895 | ClinicalTrials.gov<br>N = 1,448 | DRKS<br>N = 447 |
| --- | --- | --- | --- |
| **Time from trial completion to publication (days), Median (IQR)** | 643 (336, 1,015) | 644 (323, 1,016) | 643 (364, 1,013) |
| **Time from trial completion to summary results (days), Median (IQR)** | 536 (333, 1,017) | 544 (338, 1,028) | 344 (241, 407) |
| Unknown | 1,738 | 1,296 | 442 |
| **Registry summary results available, n (%)** | 157 (8%) | 152 (10%) | 5 (1%) |
| **Prospective registration, n (%)** | 882 (47%) | 688 (48%) | 194 (43%) |
| Unknown | 2 | 2 | 0 |
| **Randomized, n (%)** | 1,335 (84%) | 1,018 (89%) | 317 (71%) |
| Unknown | 298 | 298 | 0 |
| **Multicentric trial, n (%)** | 654 (35%) | 551 (38%) | 103 (23%) |
| Unknown | 1 | 0 | 1 |
| **Industry sponsor, n (%)** | 273 (14%) | 246 (17%) | 27 (6%) |
| **Trial enrollment, Median (IQR)** | 69 (33, 156) | 70 (34, 160) | 60 (31, 150) |
| Unknown | 4 | 4 | 0 |
| **Phase, n (%)** |  |  |  |
| I | 104 (13%) | 86 (12%) | 18 (26%) |
| I-II | 59 (7%) | 57 (8%) | 2 (3%) |
| II | 259 (32%) | 242 (32%) | 17 (24%) |
| II-III | 35 (4%) | 34 (5%) | 1 (1%) |
| III | 184 (23%) | 169 (23%) | 15 (21%) |
| IV | 175 (21%) | 158 (21%) | 17 (24%) |
| Unknown | 1,079 | 702 | 377 |
| **Top 6 journals, n (%)** |  |  |  |
| PloS One | 72 (41%) | 55 (45%) | 17 (32%) |
| Deutsches Arzteblatt International | 24 (14%) | 5 (4%) | 19 (36%) |
| BMC Cancer | 21 (12%) | 16 (13%) | 5 (9%) |
| BMC Anesthesiology | 19 (11%) | 11 (9%) | 8 (15%) |
| Lancet | 19 (11%) | 15 (12%) | 4 (8%) |
| The New England Journal of Medicine | 19 (11%) | 19 (16%) | 0 (0%) |
| **Trial completion date, n (%)** |  |  |  |
| 2009 | 107 (6%) | 97 (7%) | 10 (2%) |
| 2010 | 163 (9%) | 131 (9%) | 32 (7%) |
| 2011 | 191 (10%) | 156 (11%) | 35 (8%) |
| 2012 | 203 (11%) | 161 (11%) | 42 (9%) |
| 2013 | 195 (10%) | 160 (11%) | 35 (8%) |
| 2014 | 250 (13%) | 183 (13%) | 67 (15%) |
| 2015 | 287 (15%) | 208 (14%) | 79 (18%) |
| 2016 | 266 (14%) | 179 (12%) | 87 (19%) |
| 2017 | 233 (12%) | 173 (12%) | 60 (13%) |

Per our inclusion criteria all trials in our sample (n = 1895) were registered and had a publication, however only 373 (20%) trials had the most comprehensive registration-publication linking, meaning the publication linked in the registration as well as the TRN in the publication full-text, abstract, and PubMed metadata. Disregarding metadata, which is largely beyond trialists’ control, an additional 12 trials had comprehensive linking for a total of 385 (20%). An additional 92 (4.9%) met the CONSORT and ICMJE guidelines to include TRNs in both the full-text and the abstract. In contrast, we found 327 (17%) trials with no links in either the registration or the publication. The most common linking practice was reporting of the TRN in the full-text only, accounting for 476 (25%) trials. The inclusion of the various link types ranged from 715 (38%) in abstracts to 1,137 (60%) in full-text. Table 2 shows registration-publication links overall and by registry. Figure 1 shows the intersection of registration-publication links across the various types for ClinicalTrial.gov and DRKS trials.

**Table 2.** Registration-publication links overall and by registry.

|  | Overall<br>N = 1,895 | ClinicalTrials.gov<br>N = 1,448 | DRKS<br>N = 447 |
| --- | --- | --- | --- |
| **TRN in full-text** | 1,137 (60%) | 882 (61%) | 255 (57%) |
| **TRN in abstract** | 715 (38%) | 581 (40%) | 134 (30%) |
| **TRN in PubMed metadata** | 826 (44%) | 759 (52%) | 67 (15%) |
| **Publication in registration** | 946 (50%) | 849 (59%) | 97 (22%) |

**Figure 1.**
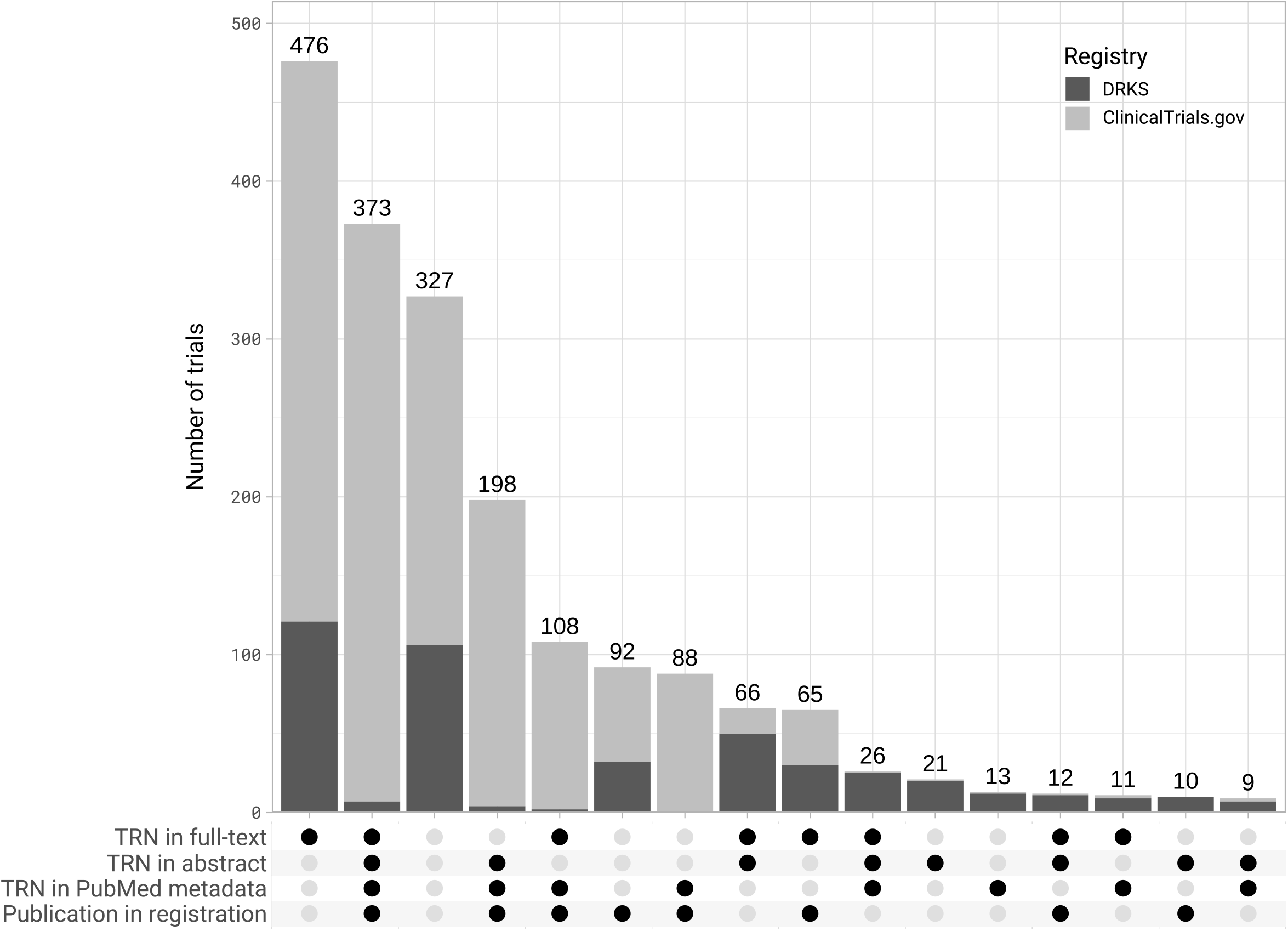
Trials with all combinations of links between registration and publication in ClinicalTrials.gov and DRKS.

Table 3 shows the crude univariate and adjusted multivariate odds ratios (ORs) for each type of publication-registration link across all explanatory variables. Completion year did not have a strong relationship with linking practices, although trials completed more recently were more likely to report the TRN in the abstract (aOR 1.13 [1.07, 1.19]) and full-text (aOR 1.07 [1.03, 1.12]). Figure 2 shows the rate of registration-publication links over time for ClinicalTrials.gov and DRKS.

**Table 3.** Crude (cOR) and adjusted (aOR) odds ratios for factors associated with publication-registration links. Odds ratio (95% CI; p value)

|  | TRN in full-text |  | TRN in abstract |  | TRN in PubMed metadata |  | Publication in registration |  |
| --- | --- | --- | --- | --- | --- | --- | --- | --- |
|  | cOR | aOR | cOR | aOR | cOR | aOR | cOR | aOR |
| **Completion year** | 1.08 (1.04, 1.12; <0.001) | 1.07 (1.03, 1.12; <0.001) | 1.07 (1.03, 1.11; <0.001) | 1.13 (1.07, 1.19; <0.001) | 0.98 (0.94, 1.02; 0.3) | 0.97 (0.91, 1.04; 0.4) | 0.96 (0.92, 0.99; 0.017) | 0.95 (0.89, 1.00; 0.067) |
| **Registry** |  |  |  |  |  |  |  |  |
| ClinicalTrials.gov | 1 (Ref) | 1 (Ref) | 1 (Ref) | 1 (Ref) | 1 (Ref) | 1 (Ref) | 1 (Ref) | 1 (Ref) |
| DRKS | 0.85 (0.69, 1.06; 0.15) | 0.76 (0.60, 0.96; 0.020) | 0.64 (0.51, 0.80; <0.001) | 4.56 (3.13, 6.75; <0.001) | 0.16 (0.12, 0.21; <0.001) | 0.22 (0.14, 0.33; <0.001) | 0.20 (0.15, 0.25; <0.001) | 0.39 (0.27, 0.55; <0.001) |
| **TRN in full-text** | NA | NA | 1.58 (1.30, 1.92; <0.001) | 1.76 (1.36, 2.28; <0.001) | 1.22 (1.02, 1.47; 0.034) | 1.33 (0.95, 1.87; 0.10) | 0.92 (0.76, 1.10; 0.4) | 0.54 (0.40, 0.72; <0.001) |
| **TRN in abstract** | 1.58 (1.30, 1.92; <0.001) | 1.78 (1.37, 2.30; <0.001) | NA | NA | 24.3 (18.9, 31.3; <0.001) | 24.2 (16.2, 37.1; <0.001) | 11.4 (9.06, 14.4; <0.001) | 2.37 (1.59, 3.49; <0.001) |
| **TRN in PubMed metadata** | 1.22 (1.02, 1.47; 0.034) | 1.25 (0.90, 1.74; 0.2) | 24.3 (18.9, 31.3; <0.001) | 26.7 (18.0, 40.8; <0.001) | NA | NA | 64.6 (47.8, 88.9; <0.001) | 37.4 (26.0, 54.9; <0.001) |
| **Publication in registration** | 0.92 (0.76, 1.10; 0.4) | 0.55 (0.42, 0.74; <0.001) | 11.4 (9.06, 14.4; <0.001) | 2.57 (1.72, 3.80; <0.001) | 64.6 (47.8, 88.9; <0.001) | 34.5 (24.0, 50.6; <0.001) | NA | NA |

**Figure 2.**
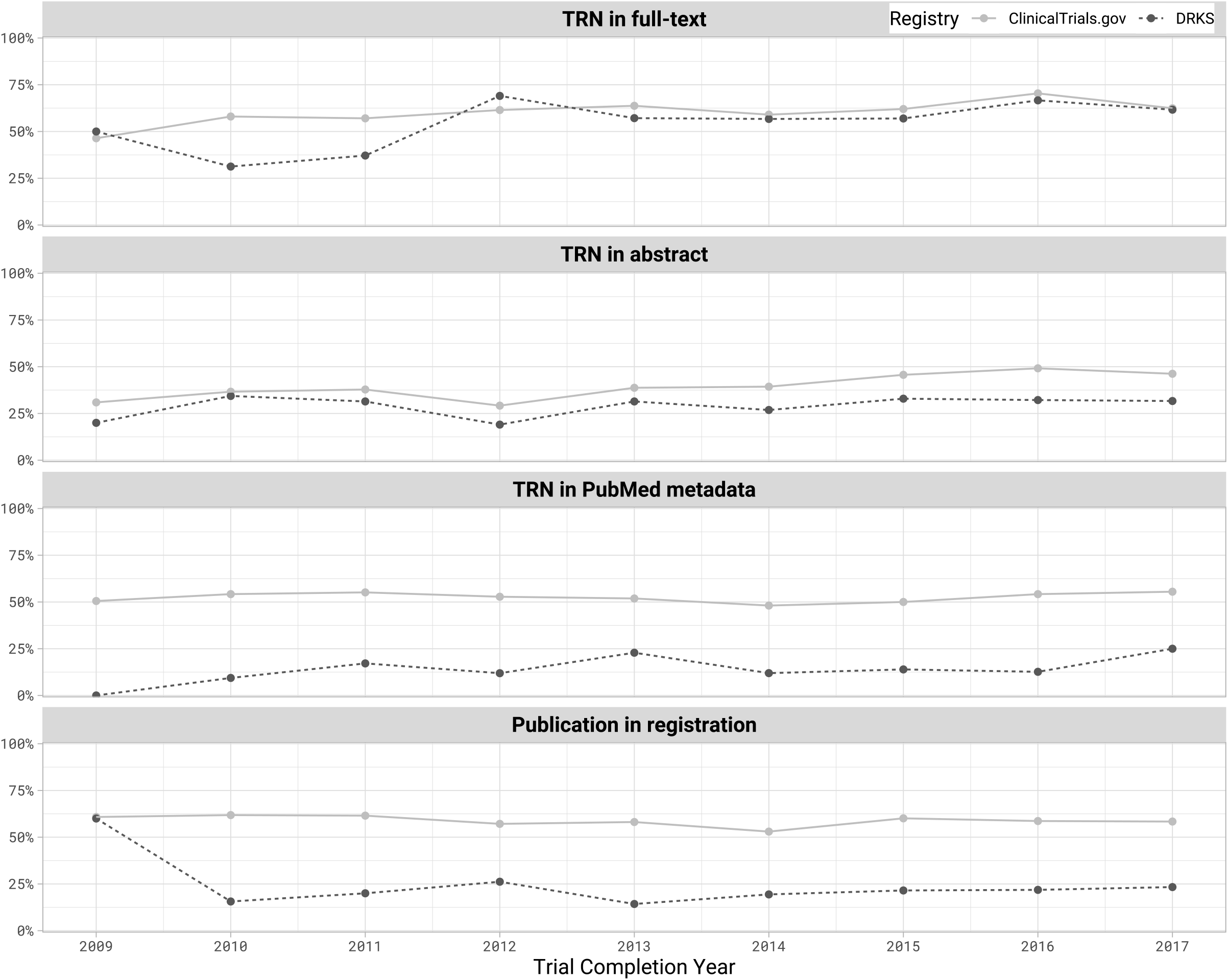
Percentage of trials with linked registrations and publications by trial completion year in ClinicalTrials.gov and DRKS. Completion year from registry.

Across the adjusted multivariate models, trials with one link type were generally more likely to have another type of link. In particular, TRNs were more likely to appear in the PubMed metadata if included in the abstract (aOR 24.2 [16.2, 37.1]), but not more likely if included in the full-text (aOR 1.33 [0.95, 1.87]). DRKS TRNs were less likely than ClinicalTrials.gov TRNs to appear in the PubMed metadata (aOR 0.22 [0.14, 0.33]). Similarly, for trials with a TRN in either abstract or full-text, ClinicalTrials.gov TRNs appeared at a higher rate than DRKS TRNs in the metadata (62% vs. 18%). Appendix B shows the number and proportion of trials with a TRN in the metadata given a TRN in the abstract, full-text, or either, both overall and by registry.

Trials registered in DRKS were also less likely than trials registered in ClinicalTrials.gov to reference the primary outcome publication in the registration (aOR 0.39 [0.27, 0.55]). Most (78%, 662/849) ClinicalTrials.gov trials that reference the publication in the registration had the link automatically derived from the PubMed metadata, which is not currently possible for DRKS. Excluding trials with automatically linked publications, ClinicalTrials.gov and DRKS had similar rates of referencing publication in the registration (24%, 187/786 vs. 22%, 97/447). Trials also had additional publications linked in the registrations, up to 72 in ClinicalTrials.gov (NCT00646412) and 13 in DRKS (DRKS00007770).

In our manual evaluation of the TRN regular expressions for classifying PubMed secondary identifiers (n = 1296), we found that most ids were clear true positive TRNs (n = 1288). A handful (n = 3) were clear true negative other ids, meaning ids from non-registry databanks, such as molecular sequences and open data repositories (i.e., figshare, Dryad) (National Library of Medicine, 2018). We also found 5 ids which we manually classified as true negative non-TRNs; however, using additional PubMed data, we determined these were severely misformatted DRKS ids (i.e., missing the preceding letters “DRKS” and just a string of numbers such as “00000711”). Our regexes correctly classified all well-formatted TRNs and non-TRNs, resulting in a sensitivity and specificity of 100%. If we had instead categorized the 5 severely misformatted DRKS ids as true positives, our regexes would have sensitivity of 99.6% and a specificity of 100%.

## 4 Discussion

Linking of trial registrations and results publications plays an important role in research transparency and facilitates comprehensive evidence synthesis and informed health policy decision making. Poor linking poses a barrier to identifying trial publications via automated approaches and instead requires researchers to perform intensive manual searches to attempt to match publications to trials (Bashir et al., 2017; Chen et al., 2016; Wieschowski et al., 2019). This responsible research practice comes at minimal costs to researchers, from seconds for pasting TRNs in papers, to minutes for adding a publication link to the registration.

Our study shows that German UMCs can improve in both TRN reporting in publications and references to publications in the registration. In our sample (n = 1895), 17% (327) of trials had no links between registration and publication and only 20% (373) of trials had the most comprehensive registration-publication links. Furthermore, only 25% (477) of trials in our sample fully met the CONSORT and ICMJE guidelines to include TRNs in both the full-text and the abstract. Linking practices showed at best minimal improvement over time. The upward trend in reporting in full-text and abstracts suggests this practice is gaining traction, however more slowly than advisable per CONSORT and ICMJE guidelines. Trials registered in ClinicalTrials.gov were overall better linked than trials registered in DRKS. These differences are in part beyond trialists’ control and reliant on bibliometric databases (i.e., PubMed) and registries, namely (1) generating PubMed metadata from TRNs in the abstract or full-text, and (2) automated indexing of publications in the registry.

Our findings suggest that PubMed’s current approach to capturing TRNs in metadata misses TRNs from the full-text as well as DRKS trials. TRNs in PubMed metadata may be either provided by publishers or manually assigned by National Library of Medicine (NLM) staff who copy-and-paste TRNs found in the abstract and full-text (personal communication via NLM Helpdesk Ticket CAS-552810-T3H7V5). As such, we expect a TRN from any registry to be included in the metadata, if the trialist includes it in either the abstract or the full-text. However, we found that while trials with a TRN in the abstract were indeed more likely to include the TRN in the metadata (aOR 24.2 [16.2, 37.1]), TRNs in the full-text were not more likely to appear in the metadata (aOR 1.33 [0.95, 1.87]). Furthermore, our data suggest that NLM staff are better at extracting ClinicalTrials.gov than DRKS TRNs (aOR 0.22 [0.14, 0.33]).

Automated indexing of publications in ClinicalTrials.gov accounts for most (78%, 662/849) publication references in the registry and drives the almost three-fold discrepancy with DRKS (59%, 849/1,448 vs. 22%, 97/447). Currently, DRKS allows for only manual submission of references by trialists and does not index publications. Furthermore, registrations may refer to additional publications beyond results (e.g., up to 72 publications for one ClinicalTrials.gov registration in our sample), so registry metadata should encode publication type to support quick identification of results. While such publication type metadata is currently available in both registries, it is not systematically used, and many publications are categorized generically as a “paper” (DRKS) or “reference” (ClinicalTrials.gov).

Accurate TRN formatting in publication data is critical for machine-readability, which in turn enables automated indexing of publications in the registration. While our regular expressions allow for and correct minor formatting errors (such as erroneous punctuation), egregious misformatting may make the TRN undetectable. For example, in our visual inspection of the PubMed metadata TRNs, we found 5 severely misformatted DRKS TRNS (i.e., numbers only with no preceding letters), which we could only identify as DRKS TRNs using additional metadata and which prevented the regex from classifying them as TRNs based on the pattern alone.

### 4.1 Strengths and Limitations

This approach has numerous strengths. In contrast with previous studies relying on PubMed queries to identify potential (randomized) clinical trial results publications, we relied on a sample of bona fide results publications from registered trials, allowing us to evaluate the rate of structured links to known results publications in its registration, and the rate of reporting a trial’s known TRN in its publication full-text, abstract, and metadata. Furthermore, we used an automated approach including regular expressions with high sensitivity and specificity to identify and classify publication identifiers and TRNs which allowed for a larger sample size that manual data extraction would have permitted. This automated strategy is scalable and can be applied to other trial sets.

The approach also faces limitations. Our input IntoValue dataset comprised German UMC-led trials in ClinicalTrials.gov and DRKS and may not reflect practices across other registries and/or countries. Future projects should look at multinational samples of bona fide results publications. Furthermore, we relied on IntoValue for trial deduplication and bona fide results publications. The dataset may have had a small number of unaccounted for cross-registrations (e.g., DRKS00004156 and NCT00215683) and publications that are not trial results (e.g., systematic reviews, conference abstract books, etc.). Finally, this automated approach faces limitations of the software on which it is built. While PubMed updated their website in 2020, their API reflects the previous backend and has subtle differences to the web version (personal communication via NLM Helpdesk Ticket CAS-575119-Y6D6Y4). While GROBID algorithms are trained on academic papers and have been used in large-scale bibliometric projects (e.g., Wang et al., 2020), parsing PDFs to XMLs may introduce some errors.

### 4.2 Implications for policy and practice

This study reveals shortcomings in the linking of German UMC-led trials registrations and results publications and highlights several promising avenues forward. In contrast with other responsible clinical research practices (such as data protection or trial registration itself), registration-publication linking is straightforward and can be improved with action across stakeholders. Table 4 outlines recommendations for stakeholders across clinical research.

**Table 4.** **Recommended stakeholder actions to improve links between trial registrations and publications**

| Stakeholder | Recommendation | Example |
| --- | --- | --- |
| **Researchers** | • Include TRN in abstract and full-text per CONSORT and ICMJE guidelines<br>• Link publication in registration | — |
| **Registries** | • Provide guidance on TRN formatting and reminder to include TRN in publications, and publications in registration<br>• Link publications automatically using TRNs in bibliographic database metadata<br>• Provide metadata on linked publication type (i.e., result, background) | ClinicalTrials.gov uses PubMed TRN metadata to automatically link publications in the registration (National Library of Medicine, 2017). Our study shows this automated publication linking is responsible for the majority of linked publications in ClinicalTrials.gov and implies similar potential for DRKS. |
| **Research Institutions** | • Educate, support, and incentivize researchers on linking | Ethics review offices or UMC-core facilities for clinical research could give researchers a info sheet with trial registration and reporting guidelines when reviewing trial. Bonuses, i.e., *Leistungsorientierte Mittel (LOM)*, could be disseminated for trial registration, reporting, and linking. |
| **Publishers/Journals** | • Request TRN in specialized metadata field<br>• Review abstract and full-text for TRN inclusion, using automated regexes and/or manual strategies<br>• Provide TRN as metadata to bibliographic databases | Taylor & Francis extracts TRNs from the abstract and full-text, submits this metadata to CrossRef and Pubmed, and displays the linked trial via Crossmark on the article page (CrossRef, 2020; Taylor & Francis Group, 2020) |
| **Bibliographic Databases** | • Integrate publisher-provided TRNs into metadata<br>• Extract TRN as metadata from abstract and full-text, using automated regexes and/or manual strategies | The National Library of Medicine relies on publisher-provided data and manual indexers to create the TRN PubMed metadata if the TRN appears in the abstract or full-text. Our study shows this manual-only strategy misses TRNs and could be semi-automated to detect more TRNs for manual verification. |

With improved TRN inclusion in bibliographic metadata and increased automatic indexing of publications based on this metadata, full linking could be achieved with negligible work by researchers: simply pasting the TRN into the publication abstract and full-text. In our sample alone, an additional 47% (650/1,375) of trials with TRN in abstract or full-text could have the TRN included in PubMed’s metadata, and an additional 79% (53/67) of DRKS trials with TRN in the PubMed metadata could be automatically indexed in the registration. By adopting the recommended actions, stakeholders can improve trial registration-publication links and foster more comprehensive evidence synthesis and well-informed clinical guidelines and health policy decisions.

## Supporting information

STROBE Checklist

## Data Availability

All code, the final analysis data, and a list of all packages used for this study are available at https://github.com/maia-sh/reg-pub-link. Raw data (with the exception of the full-text of publications) is available at https://github.com/maia-sh/intovalue-data.

https://github.com/maia-sh/intovalue-data

## 5 Acknowledgments

We would like to thank Kerstin Rubarth from the Charité Institut für Biometrie und Klinische Epidemiologie for her consultation on the statistical analysis, and Nicholas Devito for earlier versions of the trial registration number regular expressions.

## Appendix A Trial and publication screening

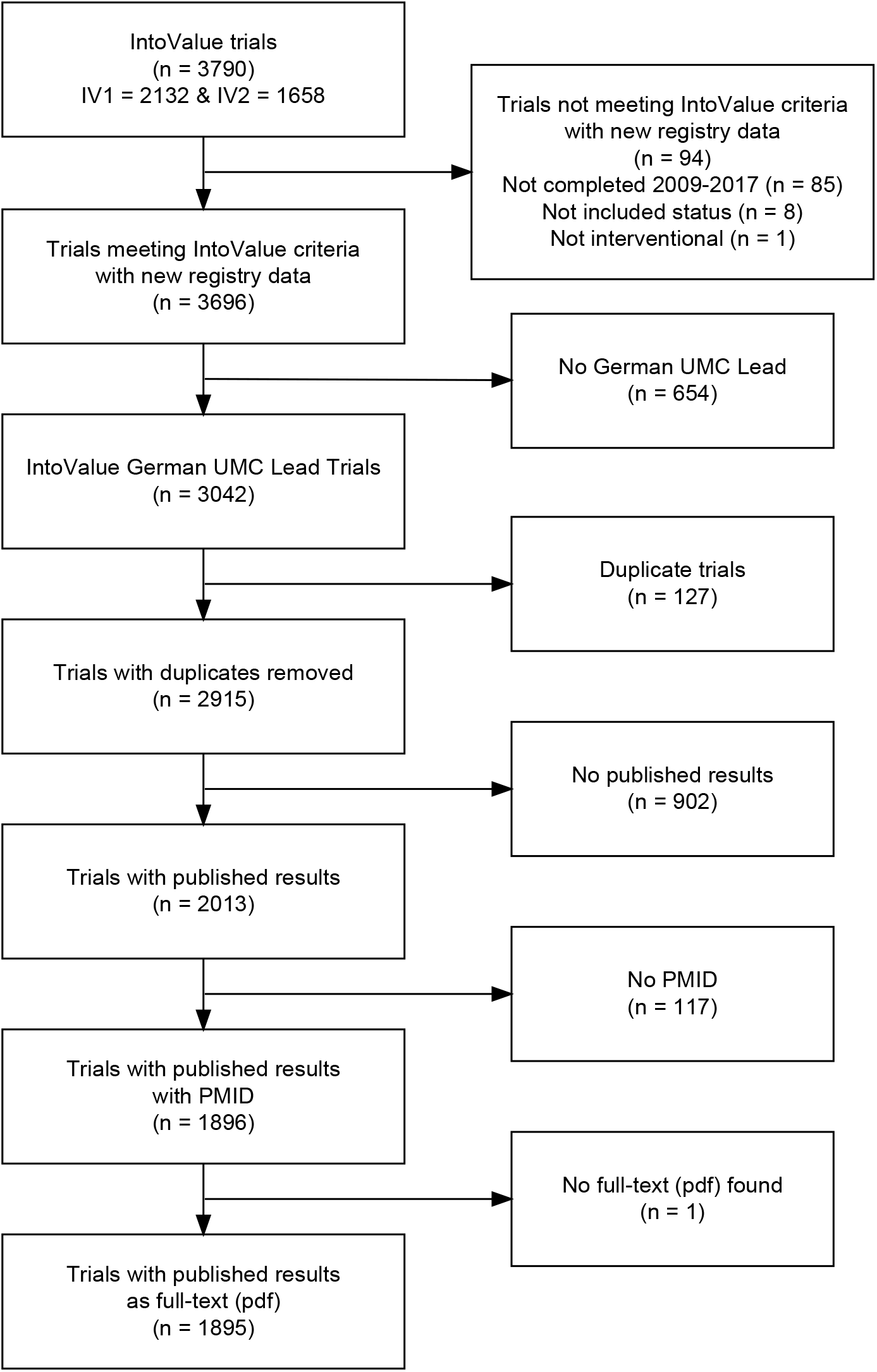

## Appendix B Number and proportion of trials with TRN in metadata given TRN in abstract, full-text, or either

Denominator in each cell indicates number of trials with TRN in abstract, full-text, or either, overall or from the respective registry.

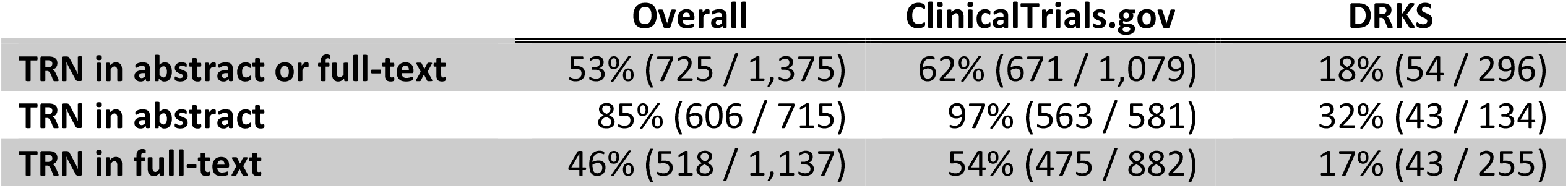

|  | Overall | ClinicalTrials.gov | DRKS |
| --- | --- | --- | --- |
| TRN in abstract or full-text | 53% (725 / 1,375) | 62% (671 / 1,079) | 18% (54 / 296) |
| TRN in abstract | 85% (606 / 715) | 97% (563 / 581) | 32% (43 / 134) |
| TRN in full-text | 46% (518 / 1,137) | 54% (475 / 882) | 17% (43 / 255) |

